# Enhancing Mental Health Condition Detection on Social Media through Multi-Task Learning

**DOI:** 10.1101/2024.02.23.24303303

**Authors:** Jiawen Liu, Menglu Su

## Abstract

**Objective:** Mental health conditions are traditionally modeled individually, which ignores the complex, interconnected nature of mental health disorders, which often share overlapping symptoms. This study aims to develop an integrated multi-task learning framework to enhance the detection of mental health conditions.

**Method:** Utilizing datasets from Reddit’s SuicideWatch and Mental Health Collection (SWMH) and Psychiatric-disorder Symptoms (PsySym), the study develops a BERT-based multi-task learning framework. This framework leverages pre-trained embedding layers of BERT variants to capture linguistic nuances relevant to various mental health conditions from social media narratives. The approach is tested against the two datasets, comparing multitask modeling with a wide array of single-task baselines and large language models (LLM).

**Results:** The multi-task learning framework demonstrated higher performance in efficiently predicting mental health conditions together compared to single-task models and general-purpose LLMs. Specifically, the framework achieved higher F1 scores across multiple conditions, with notable improvements in recall and precision metrics. This indicates more accurate modeling of mental health disorders when considered together, rather than in isolation.

**Conclusion:** The study confirms the effectiveness of a multi-task learning approach in enhancing the detection of mental health conditions from social media data. It sets a new precedent in computational psychiatry and suggests future explorations into multi-task frameworks for deeper insights into mental health disorders.

## 1. BACKGROUND

Prediction models have become an indispensable tool in the quest to diagnose and treat mental health conditions and psychiatric disorders.^1^ Historically, the focus has predominantly been on single-task modeling, where research concentrates on one specific condition at a time. This approach, while valuable, often fails to recognize the complex, interconnected nature of mental health disorders. Psychiatric disorders, as classified in diagnostic manuals such as the Diagnostic and Statistical Manual of Mental Disorders (DSM)^2^ and the International Classification of Diseases,^3^ share a wide range of symptoms that can apply across multiple conditions. For example, anxiety and depression, two of the most common mental health conditions, exhibit overlapping symptoms such as difficulty concentrating, sleep disturbances, and significant distress or impairment in social, occupational, or other important areas of functioning.^4^ This symptom overlap challenges the traditional approach of single-task modeling, which treats each condition in isolation, potentially overlooking the interconnected nature of these disorders. Ignoring the interconnected nature of these disorders can lead to a myopic understanding of mental health, potentially overlooking critical insights that could be gleaned from a more integrated approach.^**5**^

Multitask modeling has been widely studied and proved to be efficient in health care settings.^6–10^ Nonetheless, few studies have explored the area of multi-task mental disorder detection. This trend is likely due to several factors, including the methodological simplicity of single-task approaches and the historical compartmentalization of mental health research. Each condition has traditionally been studied in isolation, mirroring the structure of clinical specializations. Benton et al.^11^ paved the way with their pioneering effort to jointly model different mental disorders. Their approach utilized a straightforward feedforward neural network designed to manage multiple related tasks by sharing a single hidden layer among them. Subsequently, Buddhitha and Inkpen^12^ continued exploration of multitask mental disorder detection employing a multi-channel convolutional neural network (CNN)^13^ within a multi-task learning framework, incorporating specific auxiliary inputs to identify individuals with post-traumatic stress disorder (PTSD) and depression. Despite these methods showing potential in the collective modeling of mental disorders, they struggled to achieve good performance.

Acknowledging the valuable insights gained from previous research and recognizing their limitations, coupled with the notable advancements in Natural Language Processing (NLP), our study endeavors to develop an integrated multi-task learning framework that capitalizes on the capabilities of the Bidirectional Encoder Representations from Transformers (BERT) architecture,^14^ a landmark of the pre-training and fine-tuning era of the NLP field.^15,16^ This framework will leverage the intricate pre-trained embedding layers of BERT to simultaneously capture linguistic nuances relevant to various mental health conditions.

Our primary focus is centered on analyzing social media posts, particularly those from platforms like X (formerly known as Twitter)^17^ and Reddit.^18^ This choice allows us to gain a unique insight into the personal experiences and stories of individuals navigating the challenges of mental health, as well as participating in discussions related to these topics.^19,20^ In contrast to more restricted clinical datasets, social media data provides us with a vast and easily accessible source of information, laying a strong foundation for our initial exploration into the field of mental health research.^21,22^ The increasing trend of individuals using these platforms to openly discuss mental health issues further solidifies our choice. This trend highlights the acceptance and widespread popularity of social media data^21–27^, making it a valuable asset for our study. It ensures that our dataset is not only diverse but also reflects the complex realities of mental disorders in the real world. Additionally, the wealth of existing mental health research conducted using social media data confirms its suitability and relevance for our study.

## 2. MATERIALS AND METHODS

### 2.1. Data sources

Our research framework utilizes the Reddit SuicideWatch and Mental Health Collection (SWMH)^28^ and Psychiatric-disorder Symptoms (PsySym) datasets,^29^ informed a comprehensive review study on the application of large language models in mental health care.^20^ This review highlighted datasets covering multiple conditions. Due to recent data access restrictions by social media platforms like X and Reddit, only these two datasets were accessible for our study.

SWMH is a dataset compiled from mental health-related subreddits on Reddit, focusing on discussions around suicide intentions and mental disorders like depression and anxiety, with over 54,412 posts collected through Reddit’s API and web spiders. The dataset, aimed at identifying discussions on suicidality and mental health issues, is divided into training, validation, and testing splits, labeled by subreddit names. PsySym is another Reddit-based dataset, annotated for identifying symptoms of seven mental disorders as per DSM-5, containing 8,554 sentences across 38 symptom classes. It was meticulously annotated, involving mental health professionals, and divided into dataset splits, excluding over 40,000 control points for symptom comparison in model training and evaluation due to the study’s focus on modeling multiple disorders concurrently. Statistics of the two datasets are shown in Table 1.

**Table 1.** Distribution of Posts in SWMH and PsySym Datasets for Mental Health Analysis. Labels in SWMH were generated form post subreddit channels. Labels in PsySym were annotated with a semi-supervised method.

| Dataset | Task | Training | Validation | Test |
| --- | --- | --- | --- | --- |
| SWMH | Depression | 11,940 (34.29%) | 3,032 (34.83%) | 3,774 (34.68%) |
|  | **SuicideWatch** | 6,550 (18.81%) | 1,614 (18.54%) | 2,018 (18.54%) |
|  | **Anxiety** | 6,136 (17.62%) | 1,508 (17.32%) | 1,911 (17.56%) |
|  | **OffMyChest** | 5,265 (15.12%) | 1,332 (15.30%) | 1,687 (15.50%) |
|  | **Bipolar** | 4,932 (14.16%) | 1,220 (14.01%) | 1,493 (13.72%) |
| **PsySym** | **Anxiety** | 1412 (33.32%) | 282 (31.86%) | 1128 (32.88%) |
|  | **Depression** | 717 (16.92%) | 144 (16.27%) | 572 (16.67%) |
|  | **PTSD<sup>1</sup>** | 635 (14.98%) | 132 (14.92%) | 517 (15.07%) |
|  | **Bipolar Disorder** | 558 (13.17%) | 121 (13.67%) | 452 (13.17%) |
|  | **Eating Disorder** | 434 (10.24%) | 100 (11.30%) | 373 (10.87%) |
|  | **ADHD<sup>2</sup>** | 262 (6.18%) | 56 (6.33%) | 210 (6.12%) |
|  | **OCD<sup>3</sup>** | 220 (5.19%) | 50 (5.65%) | 179 (5.22%) |

<sup>1</sup> Post-traumatic stress disorder.
<sup>2</sup> Attention-deficit/hyperactivity disorder.
<sup>3</sup> Obsessive compulsive disorder

**Table 2.** Performance for Jointly Modeled Mental Health Conditions on SWMH and PsySym.

| Dataset | Task | F1 | Recall | Precision | Accuracy |
| --- | --- | --- | --- | --- | --- |
| SWMH | Depression | 0.8077 | 0.809 | 0.8073 | 0.9325 |
|  | SuicideWatch | 0.7973 | 0.7709 | 0.8281 | 0.9466 |
|  | Anxiety | 0.6782 | 0.6638 | 0.6941 | 0.782 |
|  | **OffMyChest** | 0.6755 | 0.6972 | 0.6556 | 0.8756 |
|  | **Bipolar** | 0.6622 | 0.5963 | 0.7457 | 0.9059 |
| **PsySym** | **Anxiety** | 83.97% | 88.57% | 79.83% | 97.93% |
|  | **Depression** | 86.35% | 91.40% | 81.83% | 90.50% |
|  | **PTSD<sup>1</sup>** | 78.02% | 92.76% | 67.32% | 94.32% |
|  | **Bipolar Disorder** | 78.68% | 71.46% | 87.53% | 94.90% |
|  | **Eating Disorder** | 84.27% | 91.68% | 77.96% | 94.84% |
|  | **ADHD<sup>2</sup>** | 94.62% | 98.32% | 91.19% | 99.42% |
|  | **OCD<sup>3</sup>** | 77.74% | 91.61% | 67.53% | 91.26% |

<sup>1</sup> Post-traumatic stress disorder.
<sup>2</sup> Attention-deficit/hyperactivity disorder.
<sup>3</sup> Obsessive compulsive disorder

### 3.2 Multi-task modeling

#### 3.2.1. Model architectures

For our multi-task modeling, we adopted the Bidirectional Encoder Representations from Transformers (BERT)-base model as the underlying architecture. This choice was informed by baseline models (introduced in next section) used in state-of-the-art studies that effectively utilized the same dataset for mental health analysis. Using the same models and same traning strategies, we manage to compare the efficiency of jointly modeling psychiatric disorders versus modeling them separately. Figure one shows a high-level illustration of our model architectures.

The process begins with raw text input, such as social media posts, which contain discussions related to mental health. The input text is tokenized using the BERT tokenizer, which breaks down the text into smaller units, or tokens, that represent words or subwords. Two special tokens are added: [CLS], which stands at the beginning and is used as an aggregate representation for classification tasks, and [SEP], which marks the end of the input or separates different sentences.

These tokens are transformed into semantic embeddings through the BERT model, also referred to as BERT embedding layers, which contains multiple layers—typically twelve in the base models. The embeddings capture the context around each token, reflecting the nuanced use of language within mental health discussions.

Following embedding, the model channels these semantic representations into the k added linear classification layers. Each layer is linked to a specific task, such as identifying signs of depression or anxiety, and culminates with a sigmoid function. The sigmoid function outputs a probability between 0 and 1, indicating the likelihood of each mental health condition being present or absent in the text.

**Figure 1:**
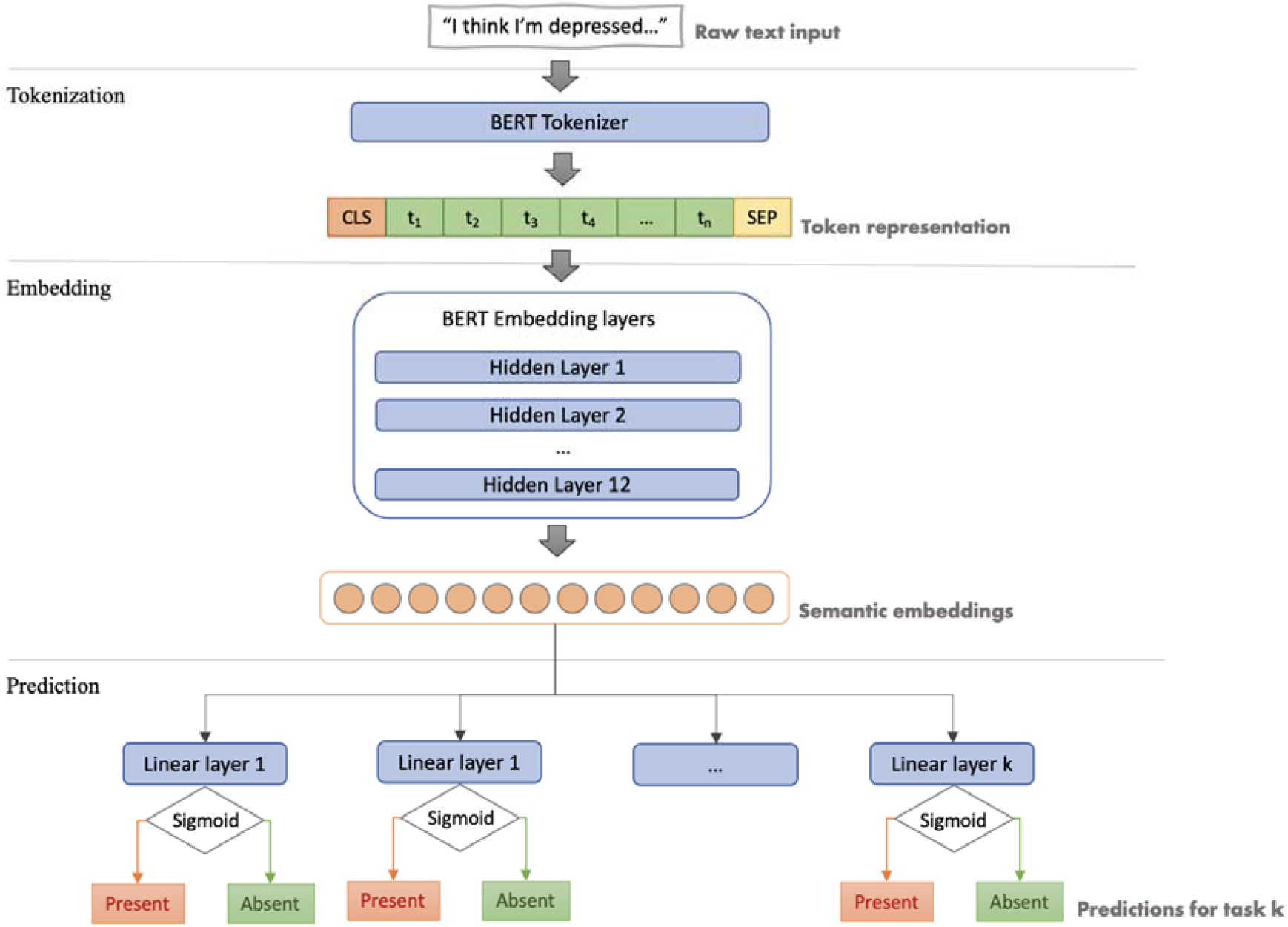
Architecture of Multi-Task Learning Models for Mental Health Condition Prediction. The figure depicts the multi-task learning model used for analyzing mental health conditions from textual data. The process starts with raw text input, followed by tokenization with the BERT tokenizer, adding special tokens such as [CLS] for classification and [SEP] for sentence separation. The tokenized text is then passed through BERT embedding layers, producing semantic embeddings. These embeddings are fed into multiple linear classification layers, each with a sigmoid function to predict the presence or absence of different mental health conditions. This model allows for simultaneous predictions across multiple tasks, enabling a comprehensive analysis of mental health-related discussions.

#### 3.2.2. Pre-trained model choices

Our selection of pre-trained models was intentionally matched with the baseline models to enable straightforward comparison. For the SWMH dataset, RoBERTa-base^30^ was identified as the best model in the paper, therefore we adopted RoBERTa as the pre-trained embedding layers for our architecture. RoBERTa is similar to the BERT model in architecture and training objectives but differs in that it is trained on a larger corpus and with longer sequences, which results in improved performance on a wide range of natural language processing tasks.

Regarding the PsySym dataset, our initial choice was BERT-base-uncased, in line with established baseline reports. However, combining modeling tasks exacerbates data scarcity, particularly when there is not a substantially balanced ratio of patients with multiple mental conditions. Consequently, we encountered challenges in training models with meaningful predictive capabilities using similar settings (e.g., learning rates and other hyperparameters) without resorting to more advanced techniques and dataset manipulations. Thus, we transitioned to RoBERTa-base, which offers similarities but boasts larger size and superior performance as pre-trained embeddings.

### 2.2. Baselines

#### 2.2.1. Single task models

Our study incorporates established baseline models referenced in prior research to benchmark and communicate our findings. Specifically, for the SWMH dataset, we scrutinize the enhancements made by Yang et al., who refined and evaluated several models, including BERT-base-uncased, RoBERTa-base, BioBERT,^31^ and ClinicalBERT.^32^ BioBERT extends the pre-training of the original BERT model to the biomedical field by utilizing research articles from PubMed.^33^ ClinicalBERT further pre-trains the BERT model using clinical notes, preparing it for enhanced performance on clinical NLP tasks. Additionally, we include MentalBERT and MentalRoBERTa, which were trained on extensive mental health datasets including SWMH.

For the PsySym dataset, we employ BERT-base-uncased as our baseline model. In addition, we explore two advanced models that leverage external information—specifically, symptom relevance—as a reference. The first model, SymP,^29^ is tailored to the task of symptom relevance judgment, with a focus on pinpointing the relevance of various symptoms to particular mental health conditions. This model signifies progress in filtering relevant symptoms from the broader textual context. The second model, SymP (Reweighting),^29^ builds upon the original SymP framework by integrating reweighting techniques to more effectively address challenges related to missing labels and data imbalance. By adjusting the weight of symptoms based on their predicted relevance and presence, this enhanced version of SymP demonstrates a sophisticated approach to navigating the intricacies of mental health symptom identification. Details of experiment settings can be found in Appendix A.

#### 2.2.2. Large Language Models

Although LLMs like GPT variants and BERT models train differently, both have become valuable tools in understanding natural language. LLMs typically learn through unsupervised learning, which means they get better at predicting text by reading lots of it. This differs from BERT’s approach, which looks at the text before and after a word to understand its meaning better.

In this study, we’re interested in how well these models can handle tasks related to mental health given their promise in general healthcare.^16,34–40^ To do this, we report several different LLMs and how they perform on two datasets. For SWMH, we report the performance of LLaMA-7B^41^ with zero prior examples given (0-shot), LLaMA-13B^41^ 0-shot, ChatGPT^42^ 0-shot, ChatGPT with a few examples given (few-shot), GPT-4^43^ few-shot, MentaLLaMA-7B^44^ few-shot, MentaLLaMA-chat-7B few-shot, and MentaLLaMA-chat-13B^7^ few-shot, published by Yang et al.^44^ For PsySym, we report the performance of ChatGPT 0-shot, as published in Chen et al.^45^

### 2.3. Evaluation

In assessing the performance of our models, we primarily employ the F1 score (binary) in alignment with the original configurations of the dataset benchmarks. To provide a more comprehensive picture of how well our models perform, we also include measurements of recall, precision, and accuracy. While we recognize that weighted metrics might be more suitable for datasets lacking balance, we have opted to adhere to the established evaluation frameworks. The absence of AUROC and AUPRC in our reporting is due to the datasets’ imbalance and the fact that the studies we are comparing our work with also did not include these metrics.

## 3. RESULTS

### 3.1. Performance Metrics on the SWMH Dataset

Upon observing the performance metrics for jointly modeled mental health conditions on the SWMH and PsySym datasets, several trends emerge. Notably, in both datasets, there is a consistent pattern of higher recall compared to precision. This indicates that the models are generally more proficient in identifying true positive cases of mental health conditions, albeit sometimes at the cost of incorrectly labeling negative cases as positive.

For the SWMH dataset, the F1 scores range from 66.22% to 80.77%, with the highest score observed for Depression and the lowest for Bipolar. Although the recall is relatively high across most tasks, precision tends to be lower, suggesting a propensity for these models to avoid false negatives at the expense of incurring more false positives. In contrast, the PsySym dataset exhibits a different pattern. Here, the F1 scores are consistently higher, spanning from 77.74% to 94.62%, with Attention-deficit/hyperactivity disorder (ADHD) showing exceptional performance across all metrics.

Interestingly, the recall rates are notably high, particularly for PTSD, ADHD, and obsessive-compulsive disorder (OCD), which have recall rates above 90%. This high recall indicates that the models are especially effective at capturing instances of these conditions. However, the precision is comparatively lower for PTSD and OCD, suggesting that while the models are adept at identifying most cases, they may also be including a number of non-relevant instances.

Accuracy across both datasets is generally high, with several conditions in PsySym showing accuracy rates above 94%. This high accuracy, especially when contrasted with the F1 scores, may suggest that the datasets are imbalanced, with a larger number of true negatives which can inflate the accuracy metric.

### 4.2. Comparison of Multi-task and Single-task Approaches

Table 4 present a comparative analysis of the performance metrics across various single-task models versus our multitask models on the SWMH and PsySym datasets. The tables illustrate how single-task models, large language models, and our multi-task framework performed in terms of F1 and Recall scores.

**Table 3.** Comparative Performance of Single-task Baseline Models and Multi-task Models on SWMH and PsySym. Color red denotes performance lower than the multitask model, color green denotes performance higher than the multitask model.

| Training Strategy | Model | SWMH |  | PsySym |
| --- | --- | --- | --- | --- |
|  |  | Recall | F1 | F1 |
| Single task | BERT | **69.78** | **70.46** | **50.8** |
|  | RoBERTa | **70.89** | **72.03** | - |
|  | BioBERT | **67.1** | **68.6** | - |
|  | ClinicalBERT | **67.05** | **68.16** | - |
|  | MentalBERT | **69.87** | **71.11** | - |
|  | MentalRoBERTa | **70.65** | **72.16** | - |
|  | Symp | - | - | **59.34** |
|  | Symp (reweighting) | - | - | **59.87** |
| Multi-task | RoBERTa | **70.74** | **72.42** | **83.37** |

**Table 4.** Performance Metrics of Large Language Models and Multi-task Framework on SWMH and PsySym Datasets. Color red denotes performance lower than the multitask model, color green denotes performance higher than the multitask model.

| Model | SWMH | PsySym |
| --- | --- | --- |
|  | F1 | F1 |
| LLaMA-7b (0 shot) | 37.33 | - |
| LLaMA-13b (0 shot) | 40.5 | - |
| ChatGPT (0 shot) | 49.32 | 60.68 |
| ChatGPT (few shot) | 60.19 | - |
| GPT-4 (few shot) | 62.94 | - |
| MentaLLaMA-7B | 72.51 | - |
| MentaLLaMA-chat-7B | 75.58 | - |
| MentaLLaMA-chat-13B | 71.7 | - |
| RoBERTa | 72.42 | 83.37 |

The RoBERTa prediction model utilizing our multi-task framework outperforms all single-task models on both datasets, as indicated by the substantially higher Recall and F1 scores. This is particularly noteworthy in the context of the SWMH dataset, where the multi-task RoBERTa achieved an 88.61% F1 score, significantly higher than that of the single-task models. The improvement is consistent across the datasets, with the multi-task approach demonstrating a F1 score of 83.3% on the PsySym dataset, which is a considerable enhancement over the Symp model’s performance, the highest among single-task approaches for this dataset.

### 4.3. Performance Metrics on the PsySym Dataset

Table 5 shows the comparison between our multi-task models and LLMs on the SWMH and PsySym datasets. We consistently observe that our multi-task framework outperforms LLMs. For example, on the SWMH dataset, the multi-task RoBERTa’s F1 score exceeds that of the highest-performing LLM, MentaLLaMA-chat-7B, by approximately 13 percentage points. In the PsySym dataset, the multi-task RoBERTa’s F1 score of 83.3% showcases its effectiveness, even surpassing the 0-shot performance of ChatGPT. Comparing Table 4 and 5, we observe that LLMs in general did not outperform light weighted BERT models.

## 5. DISCUSSION

This research represents a pioneering effort in contrasting single-task and multi-task modeling approaches within the domain of mental health analysis. Our empirical findings confirm the heightened efficacy of multi-task models over their single-task counterparts, as evidenced by higher F1 scores (72.42% on SWMH and 83.37% on PsySym). The comparative analysis outlined in Tables 3 and 4 substantiates our hypothesis of significant symptomatic and linguistic overlap among mental health conditions, with the multi-task models outperforming all the single task baselines. This overlap is well-recognized in clinical settings and is integral to diagnostic frameworks, such as those presented in the DSM and ICD, especially within the context of social media narratives. The nuanced symptomatic convergence among psychiatric conditions is particularly salient in self-reported data, where individuals may describe their experiences in a rich, idiomatic manner reflective of multiple underlying disorders.

In Tabel 2, we noted the discrepancy between recall and precision that merits attention. High recall across the board suggests that the models are adept at flagging true positives, yet this often comes at the expense of precision, indicating a tendency to also misclassify some negative instances as positive. The notably high accuracy rates, exceeding 90% for several conditions on the PsySym dataset, confer a degree of reliability to the models’ predictions. While the high accuracy rates underscore the models’ reliability in prediction, the F1 scores, which offer a more balanced view of recall and precision, underscore the potential for enhancing precision without sacrificing recall. This aspect is crucial in mental health applications where the cost of misclassification can be substantial. It is our aim to improve precision, thereby refining the models to yield more discerning predictions and achieve a more favorable balance between the two metrics.

This pursuit aligns with the findings of Chen et al.,^45^ who report binary F1 scores around 50%, in stark contrast to AUC scores of over 90%. Such a significant gap highlights a likely skewed distribution within their dataset, potentially obfuscating the true efficacy of their model. This imbalance accentuates the importance of employing weighted metrics, such as the weighted F1 score, to gain a more comprehensive understanding of a model’s performance in imbalanced datasets. Nevertheless, for consistency with established benchmarks, we opted to report binary F1 scores. Going forward, it is imperative for future research to adeptly navigate the complexities presented by class imbalances and to employ evaluation metrics that more accurately capture the true performance of models, particularly in fields as sensitive as mental health diagnostics.^46–49^

In a comparison between our lightweight multitask models and large language models (LLMs) like ChatGPT, GPT-4, and LLaMA, our models outperform general LLMs. Exceptions are MentalLLaMA 7B and MentalLLaMA-Chat 7B, trained specifically on mental health data, achieving slightly better F1 scores. However, these gains come at the cost of significant training effort, expert data curation, and substantial resources. General LLMs, trained on broad datasets, may lack nuanced expert knowledge, and fine-tuning them requires a large amount of domain-specific data, challenging in fields with data scarcity like mental health. Notably, the larger MentalLLaMA-chat-13B model underperforms compared to our models, indicating that bigger models might not always yield better results without deep domain knowledge in the mental health domain.^50–53^ This suggests that training LLMs for mental health prediction may not be economically viable, highlighting the efficiency of simpler models in achieving competitive results.

Despite the progress achieved in our study, it is important to acknowledge its limitations. Firstly, the SWMH dataset, primarily based on subreddit labels rather than expert validation, assumes subreddit discussions accurately reflect specific mental health conditions—a premise that may not be fully accurate due to the dataset’s specificity to single conditions. This limitation challenges our hypothesis that classifiers can effectively differentiate between mental health conditions using post content. The ideal dataset, previously accessible from platform X, is now difficult to obtain due to policy changes, highlighting the need for more accessible, de-identified datasets for research. Additionally, we did not analyze linguistic patterns or symptom similarities across disorders, which could provide insights into how models discern or overlook language indicative of different conditions. Furthermore, relying on social media text data omits non-verbal cues and direct clinical interactions essential for a comprehensive understanding of mental health conditions, and the assumption that social media language accurately represents these conditions may not be entirely reliable due to the influence of various factors on self-reporting.

## 4. CONCLUSION

This research highlights the effectiveness of a multi-task learning framework in improving the prediction of mental health conditions, using shared linguistic patterns across disorders for better accuracy and clinical representation. It establishes a new standard in computational psychiatry by demonstrating the benefits of integrated models over isolated ones, pointing towards significant potential for future research in understanding mental health disorders through multi-task approaches.

## Data Availability

The datasets used in this study are available through the original studies upon request.

## AUTHOR CONTRIBUTION

Conceptualization: J.Liu

Data curation: J.Liu

Formal analysis: J.Liu

Methodology: J.Liu

Supervision: J.Liu

Validation: J.Liu

Visualization: J.Liu

Writing - original draft: J.Liu

Writing - review & editing: J.Liu

## DISCLAIMER

Generative Artificial Intelligence (GAI) and AI-assisted technologies were solely utilized to enhance the readability and linguistic quality of this manuscript. All intellectual property rights associated with this study belong to the authors.

## ACKNOWLEDGEMENTS

This research received no specific grant from any funding agency in public, commercial or not-for-profit sectors.

## DATA AVAILABILITY STATEMENT

The datasets used in this study are available through the original studies upon request. Sample code used in this study will be shared online.

## CONFLICT OF INTEREST STATEMENT

The authors have no conflicts of interest to declare relevant to the content of this article.

## SUMMARY TABLE

### What was already known on the topic

- Mental health conditions and psychiatric disorders often exhibit overlapping symptoms and share many similarities in their diagnostic criteria.
- While deep learning has been applied to identify mental health conditions using social media data, few efforts were focused on simultaneously modeling multiple mental health conditions.

### What this study added to our knowledge

- This study is pioneering in comparing the approach of jointly modeling multiple mental health conditions against treating them as separate entities.
- The joint modeling framework we proposed to collectively analyze mental health conditions is shown to enhance model performance in identifying mental health conditions.
- The performance of lightweight pre-trained language models adopting our joint modeling framework even outperformed extensively fine-tuned larger language models in mental health condition detection on the same datasets.

### APPENDICES

#### Appendix A. Experiment settings

For the RoBERTa-based model on the SWMH dataset, since information about training RoBERTa was not detailed in the works of Ji et al.^54^ and Yang et al.,^44^ we opted to follow the training strategy outlined by Ji et al. for MentalRoBERTa. This strategy specified setting the learning rate for the transformer text encoder at 1e-05 and for the classification layers at 3e-05, with a batch size of 16, and performing evaluations every 1,000 steps. We also adjusted the training regimen from the extensive 624,000 iterations to a more manageable 5 epochs for our study. For the BERT-based model on the PsySym dataset, we adhered to the authors’ guidelines, setting the learning rate at 3e-4, the maximum sequence length to 64, and the number of training epochs to 4. The remaining hyperparameters were kept consistent with those of the RoBERTa model, ensuring a uniform approach to our evaluation across both models and datasets.

Both models used the Adadm optimizer.^55^ During training, all layers of the model are fine-tuned. The embedding layers learn to extract and synthesize the commonalities across different mental health conditions, capturing shared linguistic patterns. Simultaneously, the linear classification layers learn to differentiate between the conditions, recognizing condition-specific indicators within the text. This comprehensive training process ensures the model to develops nuanced understanding of mental health discourse. The embedding layers create a generalized representation, while the classification layers bring the necessary specificity for identifying individual conditions.

## Notes

### Competing Interest Statement

The authors have declared no competing interest.

### Funding Statement

This study did not receive any funding.

